# Improving UK Biobank algorithms for Parkinson’s disease ascertainment using drug claims

**DOI:** 10.64898/2026.11.18.26368409

**Authors:** Véra Kimmerling, Eva Jaber, Cécile Proust-Lima, Thérèse Truong, Alexis Elbaz

## Abstract

**Background:** Accurate ascertainment of Parkinson’s disease (PD) is a major challenge in large-scale epidemiological studies. The UK Biobank (UKB) is widely used for PD research, but ascertainment based on hospital and death records may result in misclassification and delayed PD diagnoses.

**Objectives:** We compared different strategies for ascertaining PD in the UKB, and examined their impact on incidence and associations with risk factors and mortality.

**Methods:** We compared PD ascertained through hospital and death records (ADO-PD) to two approaches that rely on primary care data available for ∼45% of participants: diagnoses from general practitioner consultations (Clinical-PD); a novel drug-based algorithm (Drug-PD). We compared incidence, age at diagnosis, and associations with sex, walking speed, and mortality across all algorithms.

**Results:** ADO-PD identified ∼20% fewer prevalent and incident cases than Drug-PD over 15y of follow-up, and estimated age at diagnosis to be 4.2y (median) later than age at anti-PD drugs initiation. ADO-PD identified never treated participants or with other causes of parkinsonism as PD patients. While Drug-PD yielded incidence rates consistent with UK reference rates, ADO-PD underestimated incidence by 36% compared to reference rates. Associations with sex, walking speed, and mortality were significantly different for ADO-PD compared to Drug-PD. Clinical-PD and Drug-PD yielded similar findings, although Clinical-PD included patients with other causes of parkinsonism.

**Conclusions:** We draw attention to methodological issues regarding PD ascertainment in the UKB and their consequences. These results can help inform strategies for PD ascertainment by combining multiple data sources, thereby improving the validity of epidemiological analyses.

## Introduction

Parkinson’s disease (PD) ascertainment represents a significant challenge in epidemiological studies. The most accurate approach relies on direct clinical assessment by trained physicians and medical records review. This approach, often used in population-based door-to-door studies in which individuals identified through screening undergo clinical examination, is often considered the reference method as it allows to exclude other causes of parkinsonism and detect undiagnosed PD. However, it is expensive and difficult, or even impossible, to implement in large cohorts involving thousands of participants or in health administrative databases containing de-identified information. An alternative solution is to use routinely collected healthcare data (e.g., outpatient diagnoses, hospital records, prescription data, mortality registries). Each data source has inherent limitations and performs differently regarding sensitivity/specificity. Outcome misclassification arising from these sources, either alone or in combination, can bias incidence estimates and associations with risk factors, particularly when misclassification is differential. Ideally, validation studies are required to describe the extent of diagnostic misclassification, estimate the impact of misclassification, and obtain corrected estimates.^1,2^ Furthermore, the availability and performance of PD ascertainment sources can be influenced by differences in healthcare systems.

The UK Biobank (UKB) is a prominent resource widely used for PD research. It is an ongoing cohort study of ∼500,000 participants whose data are linked to electronic health administrative datasets, including hospital inpatient records and death records, used to ascertain PD over the follow-up. However, most PD diagnoses occur during specialists/GP outpatient consultations, and many patients are not hospitalized during the early disease stages.^3^ Additionally, death records fail to identify a significant proportion of patients.^4^ Two studies showed that PD ascertainment in the UKB based on hospital/death records leads to under-ascertainment and delayed diagnoses. One study used general practitioner (GP) diagnoses to show that ∼30% of incident PD were not recorded in hospital/death data after 7y of follow-up; moreover, incident PD was recorded 2.3y (interquartile range=0.8-4.6) earlier in GP than in hospital/death records.^5^ Another study showed that baseline self-reports of age at PD diagnosis were 5-7y lower than age at first hospitalization/death with PD.^6^

Given the extensive use of UKB data in PD research, we performed a comprehensive comparison of three approaches to ascertain PD in the UKB over an extended follow-up, including a novel drug-based algorithm. We compared incidence rates based on these approaches as well as associations of PD incidence with a risk factor (male sex)^7^ and a prediagnostic motor manifestation (walking speed),^8-12^ and the association between PD and mortality.

## Methods

### Study population

The UKB recruited over 502,000 volunteers aged 40-69y between 2006-2010. Participants were identified through National Health Service registers and invited to attend one of 22 assessment centres across England, Scotland, and Wales (participation rate=5.5%).^13^

At baseline, defined as the date of inclusion, participants completed questionnaires assessing sociodemographic, lifestyle, and health-related information (including PD diagnosis and diagnosis date), underwent physical examinations, and provided biological samples.

Hospital records (main reason, underlying conditions), available for the entire cohort since 1981 (Scotland), 1997 (England), and 1999 (Wales),^14^ represent the main source of health outcomes ascertainment over the follow-up. Death records are available since 2006. We used data available up to October-31-2022.

Primary care data, including drug claims and GP diagnoses, are available for ∼230,000 individuals (∼45%) up to 2016-2017 (depending on data supplier). While primary care data can extend several decades before baseline, historical coverage varies substantially between participants and data providers.^14^ In addition, the number of primary care records was low before the 1980s and increased gradually thereafter,^15^ and changes in GPs or data providers can result in interruptions in primary care data for some participants. In order to use primary care data to ascertain PD and ensure the ability to distinguish prevalent from incident patients, we retained all participants with primary care data available both after baseline and at least 1y before (excluding participants whose follow-up for primary care ended before baseline, and those whose primary care data started after baseline).

As of Oct-8-2026, PubMed retrieved 334 PD studies using UKB data (Methods S1).

The UKB was approved by the North West Research Ethics Committee (06/MRE08/65). All participants signed informed consent.

### PD ascertainment

We employed three approaches to ascertain PD. First, we used the “Algorithmically-defined health outcome” (*ADO-PD*) developed by the UKB and implemented in previous studies. ADO-PD was defined as ≥1 PD code in hospital admissions and/or death records. Hospitalizations occurring before baseline and self-reported PD at baseline were used to define prevalent cases.^5,16,17^ The earliest PD code during follow-up was used to define ADO-PD incidence date. ADO-PD was ascertained both during the period with primary care data (until 2016-2017) and after (until October-31-2022) (Figure 1).

**Figure 1.**
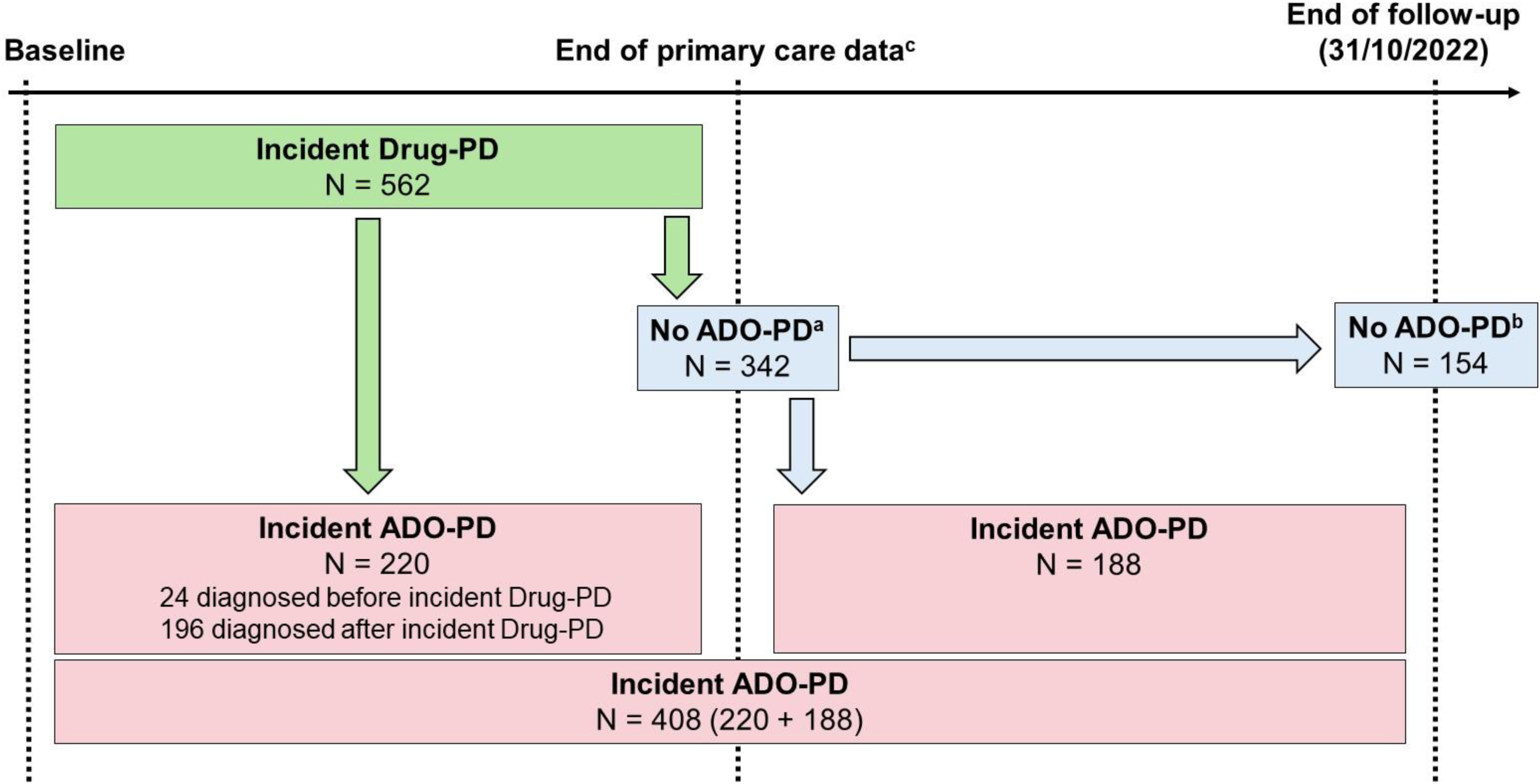
Identification over the follow-up of patients with ADO-PD in patients with incident Drug-PD. ^a^ At the end of primary care data. ^b^ At the end of the follow-up. ^c^ The maximum end of primary data is 2016-2017 depending on the data supplier. However, for individual patients, availability of primary care data can be shorter for instance if they change GP or data supplier. Between baseline and the date of end of primary care data, we identified 562 Drug-PD patients of whom 220 developed ADO-DP during the same period. Of the 342 Drug-PD patients who did not develop ADO-PD before the date of end of primary care data, 188 later developed ADO-PD between the end of the primary care data and the end of the follow-up. In total, of 562 Drug-PD patients identified before the date of end of primary care data, 408 developed ADO-PD and 154 did not develop ADO-PD before the end of the follow-up.

Second, we used GP diagnoses to define *Clinical-PD* requiring at least one PD consultation code (*Clinical-PD*) during the period with primary care data.^5^ PD patients were classified as prevalent or incident based on the earliest date of either the first GP-recorded PD diagnosis or self-reported PD.

Third, we used a sequential approach to define *Drug-PD* using drug claims recorded during the period with primary care data (Figure S1). In the first step, we selected participants with ≥1 anti-PD drug claim (levodopa, catechol-O-methyltransferase inhibitors, dopamine agonists, monoamine oxidase-B inhibitors, anticholinergics, amantadine); drugs were identified through codes^18^ and hand-searching free-text fields. In the second step, we used the same approach to identify neuroleptics, and excluded participants whose last neuroleptics claim was <5y before anti-PD drugs initiation to rule out probable drug-induced parkinsonism. We also used hospitalization codes to exclude participants hospitalized for other causes of parkinsonism or neurodegenerative disorders (Table S1). In the third step, we excluded participants with a single anti-PD prescription, as repeated prescriptions provide stronger evidence of ongoing treatment for parkinsonism than single prescriptions that may relate to alternative indications or represent errors.

We then classified anti-PD drug users as having PD based on whether their prescription profile was PD-compatible (Table S2). Dopamine agonists can be used for restless leg syndrome (RLS), generally at lower doses than for PD.^19^ Based on RLS treatment guidelines,^20^ usual doses in clinical trials, and the UK electronic medicines compendium (https://www.medicines.org.uk/emc), we considered participants receiving only low-dose dopamine agonists (rotigotine <3 mg; ropinirole <2 mg; pramipexole <0.520 mg) as more likely to have RLS. In addition, low dose bromocriptine (<5 mg) and cabergoline (<2 mg) can be used for lactation suppression. Higher doses of dopamine agonists were considered as PD-compatible. Finally, participants receiving levodopa (+/- another drug) or high-dose dopamine agonists (+/- another drug other than levodopa) were classified as having PD; users of all other combinations of anti-PD drugs were considered unlikely to have PD. In sensitivity analyses, we used a more specific definition (Levodopa-PD=levodopa +/- another drug); this approach did not rely on dopamine agonist cutoffs described above.

Drug-PD cases were classified as prevalent if the earliest date of either a first anti-PD drug claim or self-reported PD occurred before baseline, and as incident if the earliest anti-PD claim occurred after baseline.

### Covariates

We used genetic sex from available genotypes (98% of participants), and sex from a National Health Service registry or self-reported otherwise.^21^

At baseline, participants self-reported their usual walking speed through the questionnaire (“How would you describe your usual walking pace?”) as slow, steady/average, or brisk pace.

### Statistical methods

We performed several sets of complementary analyses.

First, we described PD patients characteristics based on different definitions (overall, by prevalent/incident status) and estimated the agreement (Kappa) between ADO-PD, Clinical-PD, and Drug-PD (overall, by prevalent/incident status).^22^

Second, we estimated the median delay between Drug-PD and ADO-PD diagnoses and the cumulative incidence of ADO-PD after Drug-PD, in analyses restricted to incident Drug-PD patients. We followed Drug-PD patients from their first anti-PD drug prescription until ADO-PD diagnosis, death, or October-31-2022, whichever came first. Cumulative incidence was estimated using the nonparametric Aalen–Johansen estimator, considering death as a competing risk.

Third, in the eligible population (all participants with primary care data available both after baseline and at least 1y before baseline) after excluding those with prevalent PD at baseline, we estimated PD incidence rates (/1000 person-years) by 5y-age groups over the period with primary care data using the different ascertainment methods and compared them to UK reference incidence rates through standardized incidence ratios (SIR, 95% confidence intervals [CI]). Comparing incidence rates derived from different PD ascertainment methods with reference UK rates provides an indirect means of assessing their validity. As reference rates, we used incidence rates based on primary care electronic health records from the UK Clinical Practice Research Datalink (CPRD, 2011-2015),^23^ which is considered as broadly representative of the UK population.^24^ Participants were followed from baseline until PD diagnosis, death, or end of primary care data, whichever came first.

Fourth, in the same population as for incidence rates, Cox proportional hazards regression, with age as the time-scale and delayed entry, was used to estimate the association of sex and walking speed with PD incidence through hazard ratios (HR, 95% CI) over the period with primary care data. Separate analyses were conducted for each ascertainment method. Participants were followed from baseline until PD diagnosis, death, or end of primary care data, whichever came first. We compared HRs obtained through the different ascertainment strategies by computing the relative HR (RHR=ratio of two HRs), whose CIs were obtained by bootstrapping (500 replications).

Finally, we used Cox proportional hazards regression to estimate the association between time-varying PD status and all-cause mortality over the same period; participants transitioned from PD=0 to PD=1 at the age of PD diagnosis and remained in the PD=1 state thereafter.

In sensitivity analyses, all analyses were repeated for Levodopa-PD rather than Drug-PD.

## Results

### Study sample

Of the 177,689 participants who met the inclusion criteria (Figure S1), during the period with primary care data, 946 were classified as Drug-PD (562 incident, 384 prevalent), 675 as ADO-PD (351 incident, 324 prevalent), and 856 as Clinical-PD (540 incident, 316 prevalent). Among Drug-PD patients, 83% used levodopa (+/- another drug) and 17% high-dose dopamine agonists (+/- another drug other than levodopa) (Table S2). Most incident ADO-PD patients (99%) were identified through hospitalizations.

Mean age at diagnosis was lower for incident Drug-PD (66.9y, standard deviation [SD]=6.2) than for ADO-PD (69.2y, SD=5.3; Table S3). The proportion of incident patients who died during follow-up was higher for ADO-PD (59.0%) than for Drug-PD (35.1%). Among deceased patients, the proportion with a PD code in the death certificate was lower for ADO-PD (60.3%) than Drug-PD (65.9%) and Clinical-PD (69.3%).

### Comparison of ascertainment strategies

During the period with primary care data, agreement between Drug-PD and ADO-PD was moderate for incident PD (Kappa=0.48; Table 1). Of 562 incident Drug-PD patients, 342 (61%) were not identified by ADO-PD, while 220 were identified by both algorithms (Figure 1). Additionally, 164 patients were identified by ADO-PD (40 prevalent, 124 incident) but not by Drug-PD (Table 1; no anti-PD drug, N=60; drug-induced parkinsonism, N=44; other causes of parkinsonism, N=35; 1 anti-PD drug prescription, N=3; prescription profile not PD-compatible, N=22).

**Table 1.**
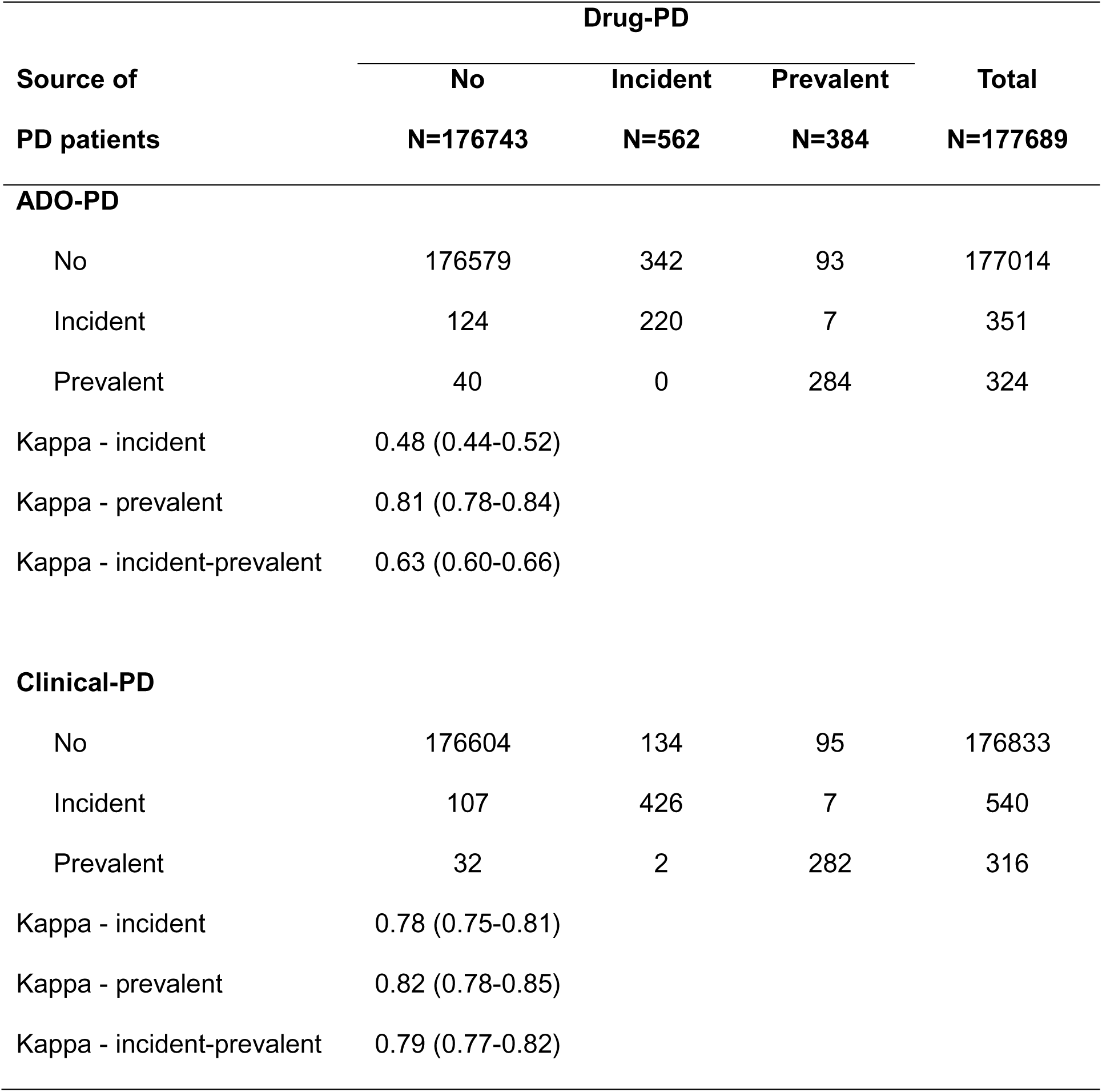
Agreement between algorithms for PD ascertainment (Drug-PD, ADO-PD, Clinical-PD) during the period with primary care data available.

| **Source of<br>PD patients** | **Drug-PD** |  |  | **Total<br>N=177689** |
|---|---|---|---|---|
|  | **No<br>N=176743** | **Incident<br>N=562** | **Prevalent<br>N=384** |  |
| **ADO-PD** |  |  |  |  |
| No | 176579 | 342 | 93 | 177014 |
| Incident | 124 | 220 | 7 | 351 |
| Prevalent | 40 | 0 | 284 | 324 |
| Kappa - incident | 0.48 (0.44-0.52) |  |  |  |
| Kappa - prevalent | 0.81 (0.78-0.84) |  |  |  |
| Kappa - incident-prevalent | 0.63 (0.60-0.66) |  |  |  |
| **Clinical-PD** |  |  |  |  |
| No | 176604 | 134 | 95 | 176833 |
| Incident | 107 | 426 | 7 | 540 |
| Prevalent | 32 | 2 | 282 | 316 |
| Kappa - incident | 0.78 (0.75-0.81) |  |  |  |
| Kappa - prevalent | 0.82 (0.78-0.85) |  |  |  |
| Kappa - incident-prevalent | 0.79 (0.77-0.82) |  |  |  |

Findings for Clinical-PD were similar to those for Drug-PD, with strong agreement for incident PD (Kappa=0.78; Table 1). However, 139 patients were identified by Clinical-PD (32 prevalent, 107 incident) but not by Drug-PD (no anti-PD drug claim, N=10; drug-induced PD, N=65; other causes of parkinsonism, N=26; 1 anti-PD drug prescription, N=9; prescription profile not PD-compatible, N=29).

Of 384 prevalent Drug-PD patients, 90 (23%) were not identified by ADO-PD by the end of follow-up and 10 (3%) were identified as incident ADO-PD patients (7 during the period with primary care data and 3 after).

To estimate the median time to ADO-PD in incident Drug-PD patients, we excluded 24 patients with ADO-PD before their first anti-PD prescription (median delay=0.4y); analyses are based on 538 incident Drug-PD patients of whom 384 developed ADO-PD before the end of the follow-up (196 during the period with primary care data, 188 between the end of primary care data and end of follow-up), 23 died before developing ADO-PD and the end of follow-up, and 131 were censored without ADO-PD at the end of the follow-up (Figure 1). The median time to ADO-PD after the first anti-PD drug prescription was 4.2y (95% CI=3.6-4.9). The 5y, 10y, and 15y cumulative incidence of ADO-PD were 54.8% (95% CI=50.5-58.9), 73.6% (95% CI=69.2-77.4), and 78.1% (95% CI=71.9-83.1) (Figure 2).

**Figure 2.**
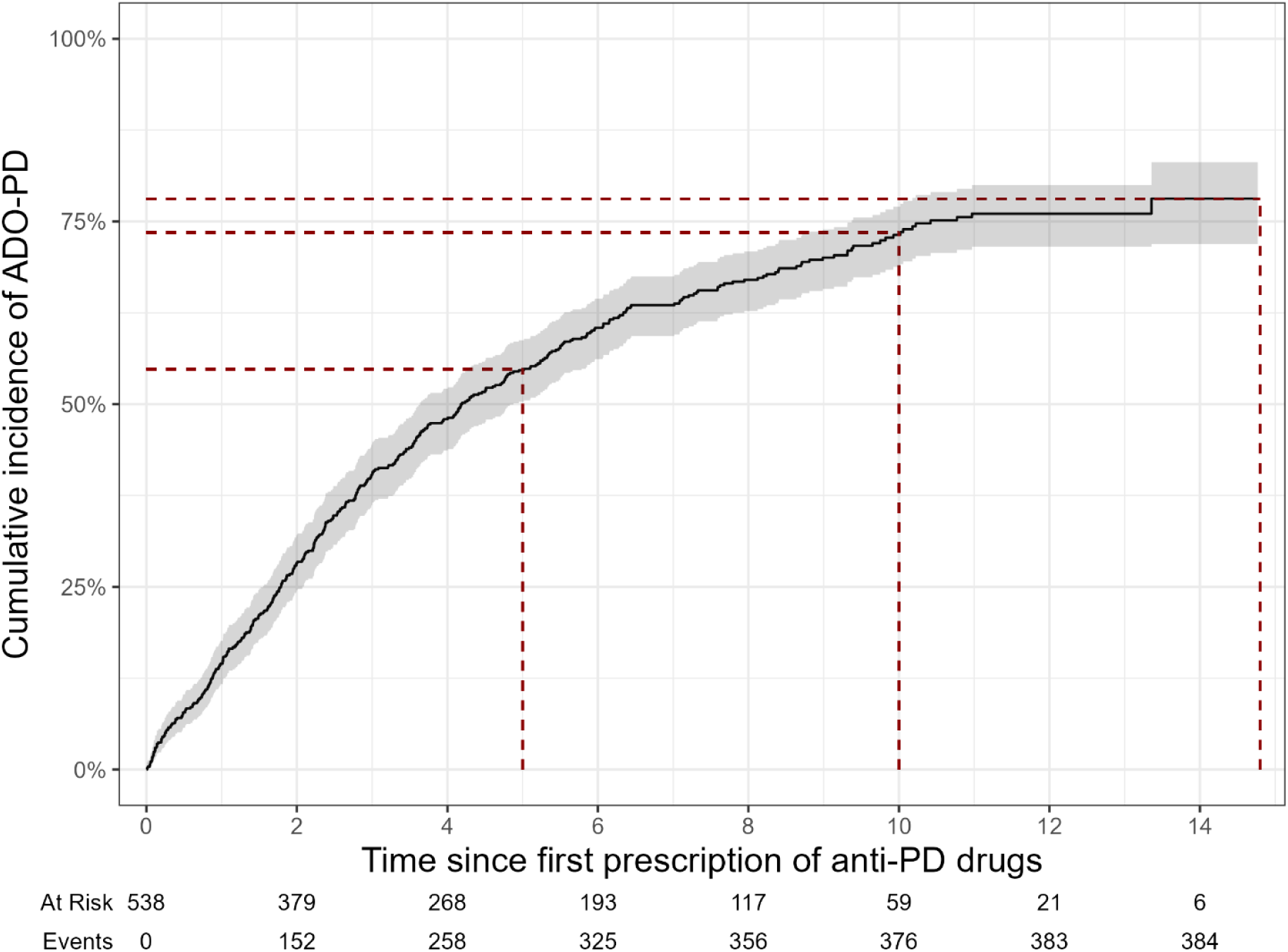
Cumulative incidence of ADO-PD in Drug-PD patients over the follow-up. Cumulative incidence (%) of ADO-PD over 15y after the first prescription of anti-PD drugs among participants identified as incident Drug-PD patients. The red dotted lines represent the 5y, 10y, and 15y cumulative incidence.

The comparison of incidence rates showed that Drug-PD (SIR=1.02, 95% CI=0.94-1.11) and Clinical-PD (SIR=0.98, 95% CI=0.90-1.06) yielded incidence rates comparable to UK reference rates, whereas ADO-PD yielded 36% lower rates (SIR=0.64, 95% CI=0.57-0.70) (Figure 3, Table S4).

**Figure 3.**
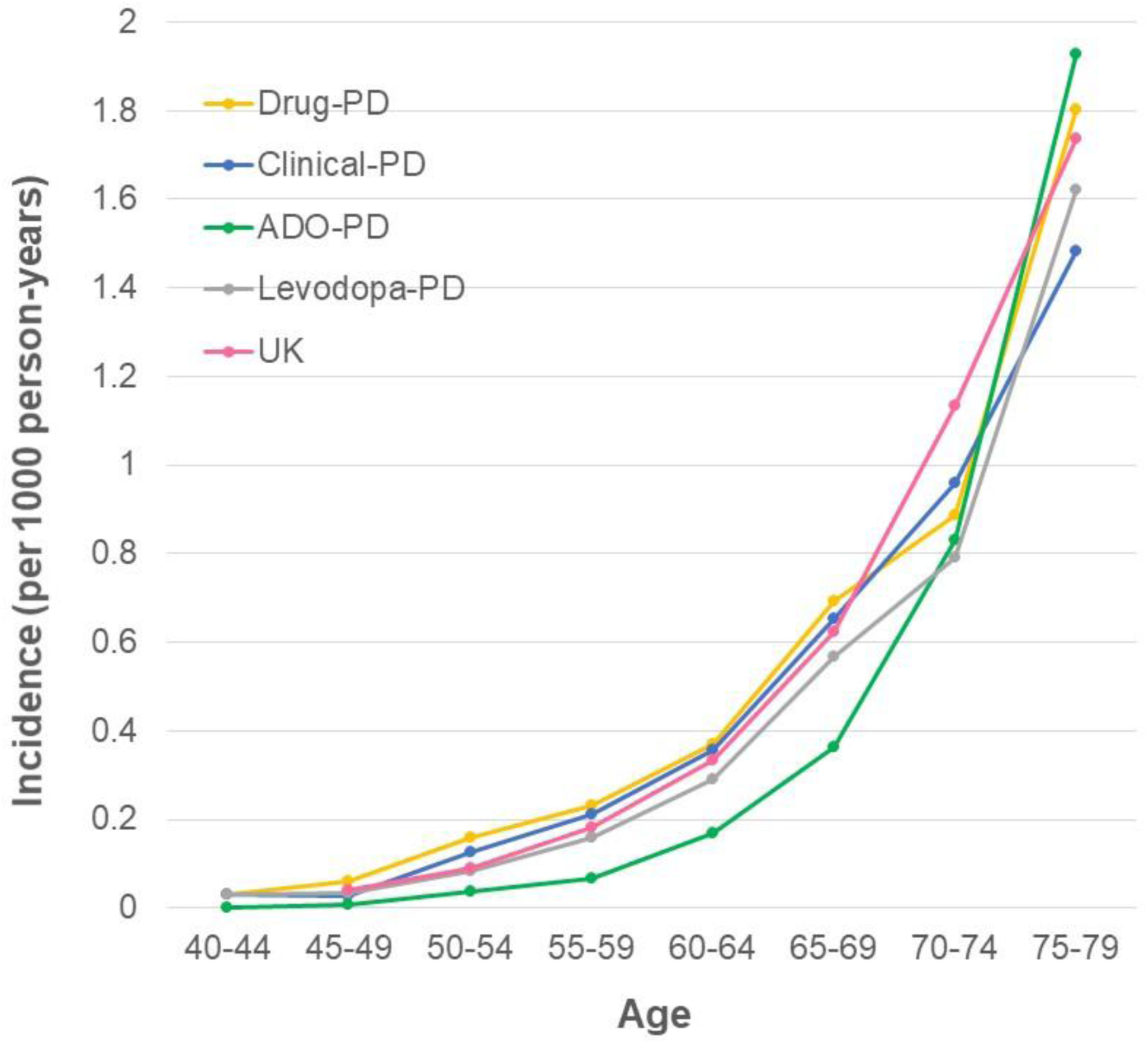
Incidence rates (per 1000 person-years) of Parkinson’s disease in the UK Biobank based on different ascertainment methods with comparison to reference incidence rates in the UK (UK Clinical Practice Research Datalink, 2011-2015). Analyses are based on the eligible population (all participants with primary care data available both after baseline and at least 1y before baseline) after excluding those with prevalent PD at baseline. Table S4 shows the number of PD patients and person-years in each age group.

### Association of sex and walking speed with PD

The association between sex and ADO-PD (HR=1.65, 95% CI=1.33-2.04) was significantly weaker than for Drug-PD (HR=2.00, 95% CI=1.68-2.37), with an estimated 17% lower HR (relative HR=0.83, 95% CI=0.71-0.98) (Table 2).

**Table 2.** Association of sex and self-reported walking speed with PD incidence over the period with primary care data.

| **Exposure** | **Level** | **PD source** | **N PD/N total** | **HR (95% CI)** | **P-value** | **Relative HR (95% CI)** |
|---|---|---|---|---|---|---|
| **Sex** | **Female** | **Drug-PD** | 209/97906 | 1.00 (reference) | -- | -- |
|  | **Male** |  | 353/79389 | 2.00 (1.68-2.37) | <10<sup>-3</sup> | -- |
|  | **Female** | **ADO-PD** | 144/97946 | 1.00 (reference) | -- | -- |
|  | **Male** |  | 207/79409 | 1.65 (1.33-2.04) | <10<sup>-3</sup> | 0.83 (0.71-0.98)<sup>a</sup> |
|  | **Female** | **Clinical-PD** | 206/97949 | 1.00 (reference) | -- | -- |
|  | **Male** |  | 334/79414 | 1.91 (1.61-2.27) | <10<sup>-3</sup> | 0.96 (0.86-1.06) |
| **Walking speed** | **Brisk** | **Drug-PD** | 148/67521 | 1.00 (reference) | -- | -- |
|  | **Slow/steady** |  | 408/108199 | 1.43 (1.18-1.72) | <10<sup>-3</sup> | -- |
|  | **Brisk** | **ADO-PD** | 68/67533 | 1.00 (reference) | -- | -- |
|  | **Slow/steady** |  | 277/108249 | 1.96 (1.50-2.56) | <10<sup>-3</sup> | 1.39 (1.12-1.71)<sup>a</sup> |
|  | **Brisk** | **Clinical-PD** | 137/67534 | 1.00 (reference) | -- | -- |
|  | **Slow/steady** |  | 398/108255 | 1.49 (1.22-1.81) | <10<sup>-3</sup> | 1.04 (0.93-1.17) |

Hazard ratios (HR) and 95% confidence intervals (CI) computed using Cox proportional hazards regression with age as the time scale; analyses for walking speed are adjusted for sex. The relative HR is computed as the HR for ADO-PD, Levodopa-PD, or Clinical-PD divided by the HR for Drug-PD; the confidence interval is obtained by the bootstrap (500 replications).
Analyses are based on the eligible population (all participants with primary care data available both after baseline and at least 1y before baseline) after excluding those with prevalent PD at baseline and with missing values for sex or walking speed.
<sup>a</sup> The 95% CI of the relative HR does not include 1.00: the difference between the HRs for ADO-PD and Drug-PD is statistically significant.

Conversely, the association between walking speed and ADO-PD (HR=1.96, 95% CI=1.50-2.56) was significantly stronger than for Drug-PD (HR=1.43, 95% CI=1.18-1.72), with an estimated 39% higher HR (relative HR=1.39, 95% CI=1.21-1.71).

Associations were similar for Drug-PD and Clinical-PD.

### Association of PD with mortality

Risk of death during the period with primary care data was higher in Drug-PD patients compared to other participants (HR=2.60, 95% CI=1.82-3.70) (Table S5). This association was 3.5-times stronger for ADO-PD (HR=8.99, 95% CI=6.79-11.89). Associations were similar for Drug-PD and Clinical-PD.

### Sensitivity analyses

Incident Levodopa-PD patients were older at PD onset than those with Drug-PD (Table S3). Except for lower incidence rates (Figure 3, Table S4), results for Levodopa-PD were comparable to those for Drug-PD (Tables S5-S7).

## Discussion

Large cohorts and health administrative databases, which generally rely on routinely collected data for PD ascertainment, are increasingly used to investigate PD risk factors. We aim to draw attention to methodological issues related to PD ascertainment and their consequences, using the UKB as a case study. We show that the UKB algorithm based on hospitalization records and death certificates (ADO-PD) (*i*) identified ∼20% fewer prevalent and incident cases than Drug-PD over 15y of follow-up, (*ii*) identified participants who were never treated or with other causes of parkinsonism as PD patients, (*iii*) estimated age at diagnosis to be a median of 4.2y later than age at anti-PD drugs initiation, (*iv*) underestimated PD incidence by 36% compared to UK reference rates, and (*v*) resulted in different associations of PD with a risk factor (sex), a prediagnostic motor manifestation (walking speed), and mortality compared to other algorithms.

PD hospitalizations were the main source of ADO-PD as most patients identified through death certificates had been previously hospitalized. PD diagnoses are predominantly made during outpatient consultations, and many patients are not hospitalized during the first years of disease. Previous research indicates that only 39% of PD patients in the UK are hospitalized during the first 5y of disease.^3^ Therefore, ADO-PD is less sensitive than Drug-PD and Clinical-PD. Age, disease severity/duration, motor fluctuations, and comorbidities are the main risk factors of hospitalizations in PD.^25^ PD patients who are hospitalized are therefore likely to have more severe PD and comorbidities than patients who are not hospitalized. In addition, patients were diagnosed with ADO-PD 4.2y later in median compared to Drug-PD, when their disease may have been more advanced. Greater severity and delayed diagnoses may explain why ADO-PD shows a considerably stronger association of ADO-PD with mortality compared to Drug-PD, Clinical-PD, and the literature.^26^

Differential diagnostic misclassification and delayed diagnoses likely explain why the association of sex and walking speed with PD varied by ascertainment strategy, as these characteristics may influence the risk of hospitalization or death in PD patients. The association between male sex and ADO-PD was weaker compared to Drug-PD and Clinical-PD. Sex may influence the risk of a first hospital encounter in PD patients^27^ and causes of hospitalization differ by sex.^3^ This could contribute to explain the weaker association of sex with ADO-PD compared to Drug-PD and Clinical-PD. In addition, men have higher all-cause mortality rates than women and a higher risk of dying from another cause than PD before they are hospitalized for PD, which could also contribute to the weaker association of male sex with ADO-PD. Regarding walking speed, several comorbidities (e.g., diabetes, hypertension, cardiovascular disease, small vessel disease) are associated with slower walking speed and higher risk of hospitalization;^25^ this may explain the stronger association between walking speed and ADO-PD compared to Drug-PD and Clinical-PD.

Delayed ADO-PD onset has important implications for risk factors analyses. When disease is identified after its true onset, exposures occurring after disease onset may be erroneously classified as preceding the disease, biasing associations with time-varying exposures.^28^ This is illustrated by a recent validation study of wearable movement tracking devices for prodromal PD detection showing that models trained on UKB data capture early clinical disease rather than prodromal markers due to delayed diagnosis and disease-stage misclassification.^29^

Even in case of unbiased associations, another consequence of incomplete case ascertainment is that it could lead to lower statistical power to detect associations and less precise association estimates.

Several previous studies used antiparkinsonian medications as “drug tracers” to ascertain PD.^19,30-33^ However, antiparkinsonian drugs may also be prescribed for other conditions. Validation studies have nevertheless shown that drug claims can be efficient when appropriate algorithms are applied. In a French study, a prediction model incorporating the type of drug, cumulative doses, and regularity of treatment achieved a sensitivity of 92.5% and specificity of 86.4% for identifying PD among antiparkinsonian drug users;^19,30^ patients on levodopa or high-dose dopamine agonists had a high probability of being treated for PD, while those on low-dose dopamine agonists were more likely to have RLS.^19^ Another study from Israel used patterns of anti-parkinsonian drugs, age at first purchase, and purchase density to develop a PD algorithm, that was highly sensitive compared to clinical diagnoses (96%).^32^ Given the lack of a similar algorithm in the UK, we used a simpler approach to ascertain PD based on drug claims. Our algorithm also allowed to rule out drug induced parkinsonism, and other causes of parkinsonism and yielded PD incidence rates consistent with those expected in the UK. Sensitivity analyses based on a more specific algorithm (Levodopa-PD) that did not rely on cutoffs for dopamine agonists showed consistent findings.

One previous study compared GP outpatient consultations to ADO-PD over a shorter follow-up and yielded findings similar to ours.^5^ In our study, results for Drug-PD and Clinical-PD also tended to be similar. However, Clinical-PD identified patients with drug-induced parkinsonism or other causes of parkinsonism as PD patients.

In this study, we specifically examined the algorithm developed by the UKB to ascertain PD. Other studies have been performed in other settings, and one systematic review included 18 studies that compared PD ascertainment using routinely collected data to a reference standard. Positive predictive value (PPV) ranged between 56-90% in hospital datasets, 53-87% in prescription datasets, and 81-90% in primary care datasets; it was 67% in mortality datasets. Combining diagnostic and medication codes increased PPV. Sensitivity ranged from 15-73% for PD.^4^ Two later US papers provide additional information. In the Rochester Epidemiology Project, compared to review of medical records by movement disorders experts, two PD International Classification of Diseases (ICD9-10) codes >30 days apart (outpatient consultations+hospitalizations) or one code in death certificates had 46% PPV and 80% sensitivity; ICD codes overestimated the date of onset on average by 1y (maximum=10y).^34^ In an extensive validation study in the Veterans Administration (VA) electronic medical records, 377 of 810 individuals with a PD ICD (9-10) code in any clinical data source (outpatient, inpatient, community care encounters records) had PD after review of their medical records by PD experts (PPV=46.5%); 98.9% were identified in outpatient data.^35^ In addition, the gold-standard diagnosis preceded the first PD ICD code by 2.5y on average. Eighteen additional PD patients were identified only from pharmacy data. The PPV increased when combining information from several sources, including provider type, number of diagnoses, and presence of dopaminergic prescriptions and exclusionary codes, although sensitivity tended to decrease.

Regarding self-report, 79.0% of participants from a French study who self-reported PD were confirmed to have PD after medical records review.^30^ In the Agricultural Health Study, 77.6% participants who self-reported physician diagnoses of PD were classified as “probable” or “possible” PD (Gelb’s criteria).^36^ In addition, while the baseline UKB assessment included the full cohort, subsequent questionnaires involved subgroups of participants; therefore, self-reports cannot be used to ascertain incident PD in the UKB.

The main strengths of this study are the comprehensive assessment of ADO-PD compared to two other algorithms, and the development of a novel drug-based algorithm to ascertain PD that we consider to be more clinically plausible.

Our study’s main limitation is that we could not validate the algorithms using PD diagnoses based on clinical examination or medical records as gold standards. We compared incidence rates derived from different PD ascertainment methods to reference UK rates as an indirect means of assessing their validity. Although agreement with reference rates supports the plausibility of an ascertainment method, this comparison does not represent a direct validation of PD identification at the individual level. In addition, the low participation rate in the UKB led to the selection of healthier participants with lower mortality rates than the general UK population.^37^ However, previous studies showed that risk factor associations with mortality in the UKB were comparable to those obtained in pooled analyses of studies with higher response rates (average, 68%).^37^ In addition, incidence rates based on Drug-PD were consistent with those estimated in the UK using primary care data, suggesting that selection effects did not significantly impact our findings. In a French cohort study of women (20% participation rate), we also found that PD incidence rates were consistent with expected rates.^30^

In conclusion, PD ascertainment in large epidemiological studies is challenging; clinical examination and medical records review are often not feasible or even impossible. PD ascertainment based on hospital records can lead to misclassification and delayed diagnoses, potentially affecting incidence estimates and associations with risk factors and mortality. Researchers using UKB data need to carefully consider the limitations described above and their potential consequences. The UKB recently announced that primary care data will be released for the whole cohort which will create an opportunity to improve PD ascertainment through anti-PD drugs and GP records. Our findings help inform strategies for combining multiple data sources to improve the validity of epidemiological analyses on PD.

## Financial Disclosure/Conflict of Interest concerning the research related to the manuscript

The authors have no financial disclosures or conflicts of interest concerning the research related to the manuscript to disclose.

## Acknowledgment

This research has been conducted using data from UKBB downloaded in November 2023 (https://www.ukbiobank.ac.uk).

## Authors’ Roles

V. Kimmerling: design, execution, analysis, writing, editing of final version of the manuscript.

E. Jaber: design, execution, editing of final version of the manuscript.

C. Proust-Lima: editing of final version of the manuscript.

T. Truong: design, editing of final version of the manuscript.

A. Elbaz: design, execution, analysis, writing, editing of final version of the manuscript.

## Financial Disclosures of all authors (for the preceding 12 months)

V. Kimmerling reports no disclosures.

E. Jaber reports no disclosures.

C. Proust-Lima reports no disclosures.

T. Truong reports no disclosures.

A. Elbaz received honorarium for speech from the Prada Foundation, and has obtained research grants from *Agence nationale de la recherche* (ANR), the Michael J Fox Foundation France Parkinson, and PEPR Prodrom.

## Data Availability Statement

This research was conducted using data from the UK Biobank under project ID 92392. Data directly supporting the results of this work are available only to the immediate research team members due to UK Biobank’s access control policy. Bona fide researchers can, however, apply for access at ukbiobank.ac.uk/enable-your-research/apply-for-access. UK Biobank data are available for approved researchers via successful application (https://www.ukbiobank.ac.uk/enable-your-research/apply-for-access).

