## Supplementary material for "Improving UK Biobank algorithms for Parkinson’s disease ascertainment using drug claims"

**Methods S1: Pubmed search of papers that used data from the UK Biobank to investigate Parkinson's disease.**……………………………………….....…...….…………………………………….…….2

**Figure S1: Flow-chart for the identification of patients with PD according to the Drug-PD algorithm.** 11

**Methods S1: Pubmed search of papers that used data from the UK Biobank to investigate Parkinson’s disease.**

We performed a PubMed search using the following terms:

("UK Biobank"[Title/Abstract] OR UKBB[Title/Abstract]) AND ("Parkinson's disease"[Title/Abstract] OR "Parkinson disease"[Title/Abstract] OR "parkinsonism"[Title/Abstract] OR "Parkinson Disease"[MeSH Terms]).

**Table S1: Exclusion of other neurodegenerative diseases.**

| | | |
|---|---|---|
| **ICD 9** | | |
| 331 | 331.0 | Alzheimer's disease |
| | 331.1 | Frontotemporal dementia |
| | 331.2 | Senile degeneration of brain |
| | 331.6 | Corticobasal Degeneration |
| | 331.7 | Cerebral degeneration in diseases classified elsewhere |
| | 331.8 | Other cerebral degeneration |
| | 331.9 | Cerebral degeneration, unspecified |
| 333 | 333.0 | Multiple System Atrophy |
| | 333.0 | PSP |
| **ICD 10** | | |
| G23 | G23.0 | Hallervorden-Spatz disease |
| | G23.1 | Progressive supranuclear ophthalmoplegia |
| | G23.2 | Striatonigral degeneration |
| | G23.3 | Hypomyelination with atrophy of basal ganglia and cerebellum |
| | G23.8 | Other specified degenerative diseases of basal ganglia |
| | G23.9 | Degenerative disease of basal ganglia, unspecified |
| G30 | G30.0 | Alzheimer disease with early onset |
| | G30.1 | Alzheimer disease with late onset |
| | G30.8 | Other Alzheimer disease |
| | G30.9 | Alzheimer disease, unspecified |
| G31 | G31.0 | Circumscribed brain atrophy |
| | G31.1 | Senile degeneration of brain, not elsewhere classified |
| | G31.2 | Degeneration of nervous system due to alcohol |
| | G31.8 | Other specified degenerative diseases of nervous system |
| | G31.9 | Degenerative disease of nervous system, unspecified |
| F00 | F00.0 | Dementia in Alzheimer disease with early onset |
| | F00.1 | Dementia in Alzheimer disease with late onset |
| | F00.2 | Dementia in Alzheimer disease, atypical or mixed type |
| | F00.9 | Dementia in Alzheimer disease, unspecified |
| F01 | F01.0 | Vascular dementia of acute onset |
| | F01.1 | Multi-infarct dementia |
| | F01.2 | Subcortical vascular dementia |
| | F01.3 | Mixed cortical and subcortical vascular dementia |
| | F01.8 | Other vascular dementia |
| | F01.9 | Vascular dementia, unspecified |
| F02 | F02.0 | Dementia in Pick disease |
| | F02.1 | Dementia in Creutzfeldt-Jakob disease |
| | F02.2 | Dementia in Huntington disease |
| | F02.3 | Dementia in Parkinson's disease |
| | F02.4 | Dementia in human immunodeficiency virus |
| | F02.8 | Dementia in other specified diseases classified elsewhere |
| F03 | F03 | Unspecified dementia |

ICD, international classification of diseases.

**Table S2: Diagnosis of Parkinson's disease (PD) according to drug profiles.**

| Drug profiles | | |
|---|---|---|
| **PD = YES (N=946)**<br>- **patients on levodopa +/- another drug (anticholinergics, COMT inhibitor, selegiline/rasagiline, amantadine);**<br>- **patients on high-dose dopamine agonists +/- another drug other than levodopa (anticholinergics, COMT inhibitor, selegiline/rasagiline, amantadine);**<br>- **anticholinergics + selegiline/rasagiline** | **N** | **%** |
| Levodopa | 261 | 27.6 |
| Levodopa + Anticholinergic | 13 | 1.4 |
| Levodopa + COMT inhibitor | 15 | 1.6 |
| Levodopa + Selegiline/rasagiline | 46 | 4.9 |
| Levodopa + Amantadine | 1 | 0.1 |
| Levodopa + Anticholinergic + Selegiline/rasagiline | 3 | 0.3 |
| Levodopa + COMT inhibitor + Selegiline/rasagiline | 11 | 1.2 |
| Levodopa + COMT inhibitor + Amantadine | 2 | 0.2 |
| Levodopa + Selegiline/rasagiline + Amantadine | 2 | 0.2 |
| Levodopa + Anticholinergic + COMT inhibitor + Selegiline/rasagiline | 2 | 0.2 |
| Levodopa + Dopamine agonist-High (+/- Low) | 143 | 15.1 |
| Levodopa + Dopamine agonist-High (+/- Low) + Selegiline/rasagiline | 84 | 8.9 |
| Levodopa + Dopamine agonist-High (+/- Low) + COMT inhibitor | 35 | 3.7 |
| Levodopa + Dopamine agonist-High (+/- Low) + Amantadine | 8 | 0.8 |
| Levodopa + Dopamine agonist-High (+/- Low) + COMT inhibitor + Selegiline/rasagiline | 54 | 5.7 |
| Levodopa + Dopamine agonist-High (+/- Low) + COMT inhibitor + Amantadine | 12 | 1.3 |
| Levodopa + Dopamine agonist-High (+/- Low) + Selegiline/rasagiline + Amantadine | 20 | 2.1 |
| Levodopa + Dopamine agonist-High (+/- Low) + COMT inhibitor + Selegiline/rasagiline + Amantadine | 35 | 3.7 |
| Levodopa + Dopamine agonist-High (+/- Low) + Anticholinergic | 9 | 1.0 |
| Levodopa + Dopamine agonist-High (+/- Low) + Anticholinergic + COMT inhibitor | 3 | 0.3 |
| Levodopa + Dopamine agonist-High (+/- Low) + Anticholinergic + Selegiline/rasagiline | 3 | 0.3 |
| Levodopa + Dopamine agonist-High (+/- Low) + Anticholinergic + Amantadine | 2 | 0.2 |

| Drug profiles | | |
|---|---|---|
| Levodopa + Dopamine agonist-High (+/- Low) + Anticholinergic + COMT inhibitor + Selegiline/rasagiline | 7 | 0.7 |
| Levodopa + Dopamine agonist-High (+/- Low) + Anticholinergic + COMT inhibitor + Amantadine | 3 | 0.3 |
| Levodopa + Dopamine agonist-High (+/- Low) + Anticholinergic + Selegiline/rasagiline + Amantadine | 1 | 0.1 |
| Levodopa + Dopamine agonist-High (+/- Low) + Anticholinergic + COMT inhibitor + Selegiline/rasagiline + Amantadine | 13 | 1.4 |
| Dopamine agonist-High (+/- Low) | 123 | 13.0 |
| Dopamine agonist-High (+/- Low) + Anticholinergic | 2 | 0.2 |
| Dopamine agonist-High (+/- Low) + Selegiline/rasagiline | 16 | 1.7 |
| Dopamine agonist-High (+/- Low) + Amantadine | 14 | 1.5 |
| Dopamine agonist-High (+/- Low) + Anticholinergic + Selegiline/rasagiline | 1 | 0.1 |
| Dopamine agonist-High (+/- Low) + Selegiline/rasagiline + Amantadine | 2 | 0.2 |
| **PD = NO (N=1217)**<br>- **patients on low-dose dopamine agonists +/- another drug other than levodopa (anticholinergics, selegiline/rasagiline, amantadine); amantadine; apomorphine (only men);**<br>- **selegiline/rasagiline alone** | **N** | **%** |
| Dopamine agonist-Low | 670 | 55.1 |
| Dopamine agonist-Low + Anticholinergic | 7 | 0.6 |
| Dopamine agonist-Low + Anticholinergic + Selegiline/rasagiline | 1 | 0.1 |
| Dopamine agonist-Low + Amantadine | 7 | 0.6 |
| Anticholinergic | 326 | 26.8 |
| Anticholinergic + Amantadine | 1 | 0.1 |
| Apomorphine* | 93 | 7.6 |
| Selegiline/rasagiline | 23 | 1.9 |
| Amantadine | 89 | 7.3 |

* All men.

High-dose dopamine agonists: rotigotine ≥3 mg; pergolide; ropinirole ≥2.0 mg; pramipexole ≥0.520 mg; bromocriptine ≥5 mg; cabergoline ≥2 mg.

Low-dose dopamine agonists: rotigotine <3 mg; ropinirole <2 mg; pramipexole <0.520 mg; bromocriptine <5 mg; cabergoline <2 mg.

**Table S3: Characteristics of participants who developed PD during the period with primary care data available according to three different algorithms.**

| Characteristics | Drug-PD | | | Levodopa-PD | | |
|---|---|---|---|---|---|---|
| | Total N=946 | Incident N=562 | Prevalent N=384 | Total N=788 | Incident N=457 | Prevalent N=331 |
| Mean age at baseline, SD | 62.2 (6.1) | 62.6 (5.8) | 61.7 (6.4) | 62.8 (5.7) | 63.2 (5.5) | 62.2 (5.9) |
| Mean age at PD onset, SD | 63.0 (8.3) | 66.9 (6.2) | 57.3 (7.8) | 63.5 (8.0) | 67.6 (5.9) | 57.8 (7.1) |
| Mean delay between age at PD onset and baseline, SD | -- | 4.3 (2.2) | -5.3 (4.5) | -- | 4.4 (2.2) | -5.4 (4.5) |
| Sex (N, %) | | | | | | |
| Female | 372 (39.3) | 209 (37.2) | 163 (42.5) | 291 (36.9) | 158 (34.6) | 133 (40.2) |
| Male | 574 (60.7) | 353 (62.8) | 221 (57.6) | 497 (63.1) | 299 (65.4) | 198 (59.8) |
| Deaths (N, %)[a] | 355 (37.5) | 197 (35.1) | 158 (41.2) | 326 (41.4) | 180 (39.4) | 146 (44.1) |
| Mean age at death, SD | 74.5 (5.3) | 75.0 (4.7) | 73.8 (6.0) | 74.5 (5.2) | 74.9 (4.7) | 74.1 (5.7) |
| Deaths with a PD code (N, %)[b] | 234 (65.9) | 125 (63.5) | 109 (69.0) | 226 (69.3) | 120 (66.7) | 106 (72.6) |
| Mean age at death with a PD code, SD | 75.2 (4.6) | 75.3 (4.5) | 75.1 (4.8) | 75.2 (4.7) | 75.3 (4.5) | 75.1 (4.9) |
| PD self-report at baseline (N, %) | 276 (29.2) | -- | 276 (71.9) | 265 (33.6) | -- | 265 (80.1) |
| Baseline self-reported walking speed (N, %)[c] | | | | | | |
| Slow | 219 (23.7) | 72 (13.0) | 147 (40.0) | 198 (25.8) | 58 (12.8) | 140 (44.3) |
| Steady | 496 (53.7) | 336 (60.4) | 160 (43.5) | 412 (53.7) | 284 (62.8) | 128 (40.5) |
| Brisk | 209 (22.6) | 148 (26.6) | 61 (16.5) | 158 (20.1) | 110 (24.3) | 48 (15.2) |
| Missing | 22 (2.3) | 6 (1.1) | 16 (4.2) | 20 (2.5) | 5 (1.1) | 15 (4.5) |
| | **ADO-PD** | | | **Clinical-PD** | | |
| | **Total N=675** | **Incident N=351** | **Prevalent N=324** | **Total N=856** | **Incident N=540** | **Prevalent N=316** |
| Mean age at baseline, SD | 63.1 (5.4) | 63.9 (5.0) | 62.3 (5.6) | 62.7 (5.5) | 62.9 (5.6) | 62.4 (5.4) |
| Mean age at PD onset, SD | 63.5 (8.7) | 69.2 (5.3) | 57.3 (7.3) | 63.5 (8.1) | 67.2 (5.9) | 57.0 (7.1) |
| Mean delay between age at PD onset and baseline, SD | -- | 5.2 (2.0) | -5.0 (4.5) | -- | 4.4 (2.3) | -5.4 (4.6) |
| Sex (N, %) | | | | | | |
| Female | 267 (39.6) | 144 (41.0) | 123 (38.0) | 326 (38.1) | 206 (38.2) | 120 (38.0) |
| Male | 408 (60.4) | 207 (59.0) | 201 (62.0) | 530 (61.9) | 334 (61.8) | 196 (62.0) |
| Deaths (N, %)[a] | 335 (49.6) | 182 (51.9) | 153 (47.2) | 348 (40.7) | 198 (36.7) | 150 (47.4) |
| Mean age at death, SD | 74.1 (5.4) | 74.5 (4.9) | 73.5 (5.9) | 74.4 (5.2) | 75.0 (4.8) | 73.7 (5.7) |
| Deaths with a PD code (N, %)[a] | 202 (60.3) | 94 (51.6) | 108 (70.6) | 237 (68.1) | 130 (65.7) | 107 (71.3) |
| Mean age at death with a PD code, SD | 75.0 (4.6) | 75.1 (4.5) | 74.9 (4.8) | 75.1 (4.6) | 75.3 (4.5) | 75.0 (4.8) |
| PD self-report at baseline (N, %) | 305 (45.2) | -- | 305 (94.1) | 305 (35.6) | -- | 305 (96.5) |
| Baseline self-reported walking speed (N, %)[b] | | | | | | |
| Slow | 196 (30.1) | 58 (16.8) | 138 (45.1) | 196 (23.5) | 62 (11.6) | 134 (44.8) |
| Steady | 338 (51.9) | 219 (63.5) | 119 (38.9) | 453 (54.3) | 336 (62.8) | 117 (39.1) |
| Brisk | 117 (20.0) | 68 (19.7) | 49 (16.0) | 185 (22.2) | 137 (25.6) | 48 (16.1) |
| Missing | 24 (3.6) | 6 (1.7)) | 18 (5.6) | 22 (2.6) | 5 (0.9) | 17 (5.4) |

SD, standard deviation.

[a] During the whole follow-up (until October-31-2022).

[b] The percentage of patients with a PD code in death certificates is computed among those who died.

[c] The denominator used to compute the percentage of non-missing categories does not include the missing values; the denominator for the percentage of missing values is the total number of subjects.

**Table S4: Incidence rates (per 1000 person-years) of Parkinson's disease (PD) in the UK Biobank over the period with primary data based on four algorithms compared to reference incidence rates in the UK.**

| | Drug-PD | | | Levodopa-PD | | | Reference, UK |
|---|---|---|---|---|---|---|---|
| **Age** | **Person-years** | **N PD** | **Incidence** | **Person-years** | **N PD** | **Incidence** | **incidence rates[a]** |
| **40-44** | 34242.2 | 1 | 0.03 | 34238.8 | 1 | 0.03 | -- |
| **45-49** | 117075.7 | 7 | 0.06 | 117051.9 | 4 | 0.03 | 0.04 |
| **50-54** | 158746.5 | 25 | 0.16 | 158700.8 | 13 | 0.08 | 0.09 |
| **55-59** | 193167.5 | 45 | 0.23 | 193104.6 | 31 | 0.16 | 0.18 |
| **60-64** | 258081.2 | 95 | 0.37 | 257960.1 | 75 | 0.29 | 0.34 |
| **65-69** | 292222.7 | 202 | 0.69 | 292116.6 | 166 | 0.57 | 0.62 |
| **70-74** | 165503.3 | 147 | 0.89 | 165460.6 | 131 | 0.79 | 1.13 |
| **75-79** | 22204.3 | 40 | 1.80 | 22198.2 | 36 | 1.62 | 1.74 |
| **Total** | 1241243.4 | 562 | 0.45 | 1240831.6 | 457 | 0.37 | -- |
| **SIR** | 1.02 (0.94-1.11) P=0.62 | | | 0.83 (0.75-0.91) P<0.0001 | | | |

| | ADO-PD | | | Clinical-PD | | | Reference, UK |
|---|---|---|---|---|---|---|---|
| **Age** | **Person-years** | **N PD** | **Incidence** | **rates[a]** | **N PD** | **Incidence** | **incidence rates[a]** |
| **40-44** | 34260.4 | 0 | 0.00 | 34260.1 | 1 | 0.03 | -- |
| **45-49** | 117133.6 | 1 | 0.01 | 117127.8 | 3 | 0.03 | 0.04 |
| **50-54** | 158848.3 | 6 | 0.04 | 158831.3 | 20 | 0.13 | 0.09 |
| **55-59** | 193338.0 | 13 | 0.07 | 193283.6 | 41 | 0.21 | 0.18 |
| **60-64** | 258366.3 | 44 | 0.17 | 258177.9 | 92 | 0.36 | 0.34 |
| **65-69** | 292664.3 | 106 | 0.36 | 292355.6 | 191 | 0.65 | 0.62 |
| **70-74** | 165910.9 | 138 | 0.83 | 165628.7 | 159 | 0.96 | 1.13 |
| **75-79** | 22288.3 | 43 | 1.93 | 22232.8 | 33 | 1.48 | 1.74 |
| **Total** | 1242809.9 | 351 | 0.28 | 1241897.7 | 540 | 0.43 | -- |
| **SIR** | 0.64 (0.57-0.70) P<0.0001 | | | 0.98 (0.90-1.06) P=0.66 | | | |

Incidence per 1000 person-years.
SIR, standardized incidence ratio with UK incidence rates as the reference
Analyses are based on the eligible population (all participants with primary care data available both after baseline and at least 1y before baseline) after excluding those with prevalent PD at baseline.
[a] Rates from the Parkinson's UK study based on Clinical Practice Research Datalink (CPRD) data between 2011 and 2015 (Table 9 from https://www.parkinsons.org.uk/sites/default/files/2018-01/Incidence%20and%20prevalence%20report%202018.pdf).

**Table S5: Association of PD with risk of death over the period with primary data.**

| PD ascertainment | PD | N deaths/ N total | HR (95% CI) | P-value | Relative HR (95% CI) |
|---|---|---|---|---|---|
| **Drug-PD** | **No** | 4618/176741 | 1.00 (reference) | -- | -- |
| | **Yes** | 31/562 | 2.60 (1.82-3.70) | <0.001 | -- |
| **Levodopa-PD** | **No** | 4618/176741 | 1.00 (reference) | -- | -- |
| | **Yes** | 29/457 | 2.90 (2.00-4.17) | <0.001 | 1.11 (0.75-1.87) |
| **ADO-PD** | **No** | 4597/177012 | 1.00 (reference) | -- | -- |
| | **Yes** | 51/351 | 8.99 (6.79-11.89) | <0.001 | 3.45 (2.78-4.48)[a] |
| **Clinical-PD** | **No** | 4619/176831 | 1.00 (reference) | -- | -- |
| | **Yes** | 32/540 | 2.71 (1.94-3.89) | <0.001 | 1.06 (0.78-1.33) |

Hazard ratios (HR) and 95% confidence intervals (CI) computed using Cox proportional hazards regression for time-varying variables with age as the time scale and adjusted for sex. The relative HR is computed as the HR for ADO-PD Levodopa-PD, or Clinical-PD divided by the HR for Drug-PD; the confidence interval is obtained by the bootstrap (100 replications).

[a] The 95% CI of the relative HR does not include 1.00: the difference between the HRs for ADO-PD and Drug-PD is statistically significant.

**Table S6: Agreement between algorithms for PD ascertainment (Levodopa-PD, ADO-PD, Clinical-PD) over the period with primary care data.**

| | Levodopa -PD | | | |
|---|---|---|---|---|
| **Source of PD patients** | **No N=176 743** | **Incident N=457** | **Prevalent N=331** | **Total N=177 531** |
| **ADO-PD** | | | | |
| No | 176 579 | 246 | 52 | 176 877 |
| Incident | 124 | 211 | 7 | 342 |
| Prevalent | 40 | 0 | 272 | 312 |
| Kappa - incident | 0.53 (0.49-0.57) | | | |
| Kappa - prevalent | 0.86 (0.83-0.88) | | | |
| Kappa - incident-prevalent | 0.68 (0.65-0.71) | | | |
| **Clinical-PD** | | | | |
| No | 176 604 | 69 | 53 | 176 726 |
| Incident | 107 | 387 | 7 | 501 |
| Prevalent | 32 | 1 | 271 | 304 |
| Kappa - incident | 0.81 (0.79-0.84) | | | |
| Kappa - prevalent | 0.85 (0.84-0.89) | | | |
| Kappa - incident-prevalent | 0.84 (0.82-0.86) | | | |

**Table S7: Association of sex and self-reported walking speed with Levodopa-PD incidence over the period with primary care data.**

| Exposure | Level | PD source | N PD/ N total | HR (95% CI) | P-value | Relative HR (95% CI) |
|---|---|---|---|---|---|---|
| **Sex** | **Female** | **Levodopa-PD** | 158/97855 | 1.00 (reference) | -- | -- |
| | **Male** | | 299/79335 | 2.22 (1.83-2.69) | <0.001 | 1.11 (0.89-1.36) |
| **Walking speed** | **Brisk** | **Levodopa-PD** | 110/67483 | 1.00 (reference) | -- | -- |
| | **Slow/steady** | | 342/108133 | 1.58 (1.27-2.00) | <0.001 | 1.11 (0.86-1.40) |

Hazard ratios (HR) and 95% confidence intervals (CI) computed using Cox proportional hazards regression with age as the time scale and adjusted for sex. The relative HR is computed as the HR for Levodopa-PD divided by the HR for Drug-PD (Table 2); the confidence interval is obtained by the bootstrap (500 replications).

**Figure S1: Flow-chart for the identification of patients with PD according to the Drug-PD algorithm.**

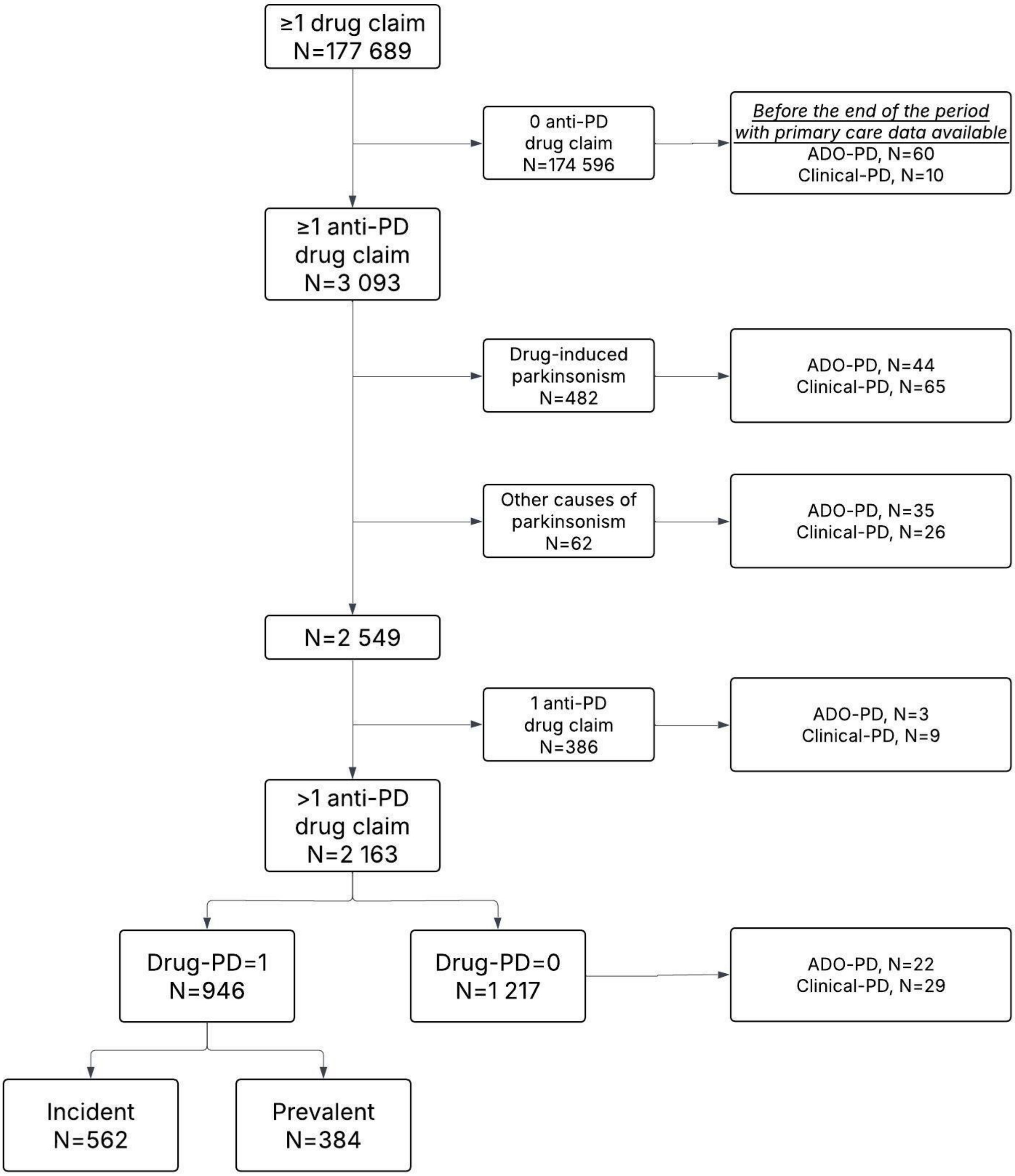

ADO-PD, algorithmically-defined health outcome based on hospitalizations and death certificates with a PD code.
Clinical-PD, PD status based on general practitioners' outpatient consultations PD codes.
